# Natural Polymorphisms in *Mycobacterium tuberculosis* Conferring Resistance to Delamanid in Drug-naïve Patients

**DOI:** 10.1101/2020.08.07.20167130

**Authors:** Martina L. Reichmuth, Rico Hoemke, Kathrin Zürcher, Peter Sander, Anchalee Avihingsanon, Jimena Collantes, Chloé Loiseau, Sonia Borrell, Miriam Reinhard, Robert J. Wilkinson, Marcel Yotebieng, Lukas Fenner, Erik C. Böttger, Sebastien Gagneux, Matthias Egger, Peter M. Keller on behalf of the International epidemiology Databases to Evaluate AIDS (IeDEA)

**Author notes:** Martina L. Reichmuth and Rico Hoemke contributed equally to this work. <u>Correspondence to</u>: Peter M. Keller, MD, Institute for Infectious Diseases, University of Bern, Friedbühlstrasse 51, 3001 Bern, Switzerland.

## Abstract

Mutations in the genes of the F_420_ signaling pathway, including *dnn, fgd1, fbiA, fbiB, fbiC*, and *fbiD*, of *Mycobacterium tuberculosis* (*Mtb*) complex can lead to delamanid resistance. We searched for such mutations among 129 *Mtb* strains from Asia, South-America, and Africa using whole-genome sequencing; 70 (54%) strains had at least one mutation in one of the genes. For ten strains with mutations, we determined the minimum inhibitory concentration (MIC) of delamanid. We found one strain from a delamanid-naïve patient carrying the natural polymorphism Tyr29del (*ddn*) that was associated with a critical MIC to delamanid.

---

In 2014, the new anti-tuberculosis (TB) drug, delamanid (also known as OPC-67683 or Deltyba™) was introduced (1). The World Health Organisation (WHO) recommends the administration of delamanid if a standard effective drug regimen cannot be prescribed due to drug toxicity or resistance (2, 3). Thus, the European Medicines Agency (EMA) conditionally approved delamanid for the treatment of multidrug-resistant (MDR) TB (1, 3, 4). Of note, six years after its market launch, robust and widely accepted breakpoints that define susceptibility and resistance to delamanid still do not exist (5). The few available studies suggest a critical MIC between 0.125 mg/L and 0.2 mg/L, and an Epidemiological Cutoff Value (ECOFF) of 0.04 mg/L (6-9). This ECOFF is in line with the WHO (10).

Delamanid is a drug of the bicyclic nitroimidazole class with potent anti-TB activity (1, 11). It is a pro-drug which is activated by the deazaflavin (F_420_) dependent nitroreductase (*ddn*) through hydride transfer, forming unstable intermediates, which in turn lead to the formation of reactive nitrogen species (nitric oxide, nitrous acid) (12, 13). Activated delamanid thus has a dual bactericidal mode of action as the primary decomposition product prevents mycolic acid synthesis while the reactive nitrogen species cause respiratory poisoning (12-15). Loss of function mutations in *ddn* or one of the five coenzymes (*fgd1, fbiA, fbiB, fbiC, and fbiD*) have been proposed as a mechanism of resistance to delamanid (12, 13, 16, 17). *In vitro*, frequencies of delamanid resistance-conferring mutations in the *Mycobacterium tuberculosis* (*Mtb*) laboratory strain *H37Rv* and in *M. bovis* range from 2.51 × 10^-5^ to 6.44 × 10^-6^ (13). Previous studies have found several resistance-conferring mutations, including Leu107Pro (ddn), 51-101del (*ddn*), Trp88STOP (*ddn*), Gly81Asp (*ddn*), Gly81Ser (*ddn*), Gly53Asp (*ddn*), c.146_147insC (*fgd1*), Gln88Glu (*fgd1*), Lys250STOP (*fbiA*), Arg175His (*fbiA*), and Val318lle (*fbC*) (6-8, 18-22).

This multicentre study has been described in detail elsewhere and is part of the International epidemiology Databases to Evaluate AIDS (IeDEA) (23). We identified putative delamanid resistance-conferring mutations in *Mtb* strains from TB patients living with HIV (PLWH) and HIV negative TB patients naïve to delamanid using whole-genome sequencing (WGS) and MIC determination. We collected demographic and clinical characteristics of patients that were recruited between 2013-2016 in Peru, Thailand, Côte d’Ivoire, Democratic Republic of the Congo (DRC), Kenya, and South Africa (24, 25). The Cantonal Ethics Committee in Bern, Switzerland, and local institutional review boards approved the study. Written informed consent was obtained at all locations, except in South Africa, where consent was not required for archived samples.

The sequencing pipeline has been described previously (25). In brief, *Mtb* DNA was extracted and sequenced using Illumina HiSeq 2500 (Illumina, San Diego, CA, USA). For the analysis, we used the well-established pipeline TBprofiler (https://github.com/jodyphelan/TBProfiler (26, 27)). It aligns short reads to the *Mtb* reference (H37Rv: NC_000962.3) with bowtie2 *v2.3.5*, BWA *v0.7.17* or minimap2 *v2.16* and then calls variants with SAMtools *v1.9* (28-31). To identify putative delamanid resistance-conferring mutations, we analysed F_420_ genes (*ddn, fgd1, fbiA, fbiB, fbiC, and fbiD*) with variant frequencies ≥75%. A subset of *Mtb* strains with at least one mutation in F_420_ genes were re-cultured in liquid medium and subjected to delamanid MIC determination (Supplementary Figure 1). We assumed that 0.04 mg/L indicates a critical MIC (9).

We included 129 *Mtb* isolates among them 52 (40.3%) from Peru, 14 (10.9%) from Thailand, 51 (39.5%) from Côte d’Ivoire, 14 (10.9%) from DRC, and 1 (0.8%) each from Kenya and South Africa. We identified 70 (54.3%) isolates with polymorphisms in at least one of the six F_420_ genes as compared to the reference genome (Supplementary Table 1). None of the patients infected with either of these strains had a history of TB and all were naïve to delamanid. We selected strains fulfilling the following criteria: i) mutations in a part of the gene encoding regions of catalytic or structural importance predicted by ARIBA and then the PhyResSE pipeline (32, 33), ii) culture of the strain available iii) bacterial growth amenable to microdilution (25). MIC determination was performed on ten isolates with mutations in the F_420_ genes. Four isolates showed a MIC >0.015 mg/L: 0.5 (patient 1), 0.03 (patients 6 and 10), and >8 mg/L (patient 9; Table 1; Supplementary Figure 1). The isolate from patient 1 had a polymorphism in *fgd1* (Lys270Met), was susceptible to the six tested drugs (isoniazid, rifampicin, ethambutol, pyrazinamide, moxifloxacin, and amikacin). The patient was cured. The isolate from patient 9 had two alterations, a deletion in *ddn* (Tyr29del) and carried a nucleotide change in *fgd1* (T960C). The strain showed an elevated delamanid MIC and was phenotypically susceptible to six other drugs tested. The patient died. Isolates of patient 10 (and 6) had a MIC above 0.015 but below 0.04 mg/L (Table 1). This suggests a low-level resistance to delamanid (22), which could be due to the combination of various mutations: Ala416Val (*fbiC*), Trp678Gly (*fbiC*), Arg64Ser (*fgd1*), and T960C (*fgd1*).

**TABLE 1.** Observed polymorphisms in F_420_ genes and minimal inhibitory concentration values for delamanid.

| No. | Lineage | Country | HIV status | Age at TB diagnosis | Gender | Mutations in the F<sub>420</sub> genes | Treatment | Treatment outcome | MIC mg/L in the micro-dilution |
| --- | --- | --- | --- | --- | --- | --- | --- | --- | --- |
| 0 | H37Rv ATCC 27294 | - | - | - | - | Control (wt) | - | - | ≤0.015 |
| 1 | L4.1.2.1 | Côte d'Ivoire | Negative | 29 | Female | *fgd1* Lys270Met | 2HRZE, 4RH | Cured | 0.5 |
| 2 | L4.6.2.2 | Côte d'Ivoire | Negative | 51 | Male | *ddn* C168T | 2HRZE, 4RH | Died | ≤0.015 |
| 3 | L2.2.1 | Kenya | Positive | 40 | Male | *fgd1* T960C | 2HRZE, 4RH | Died | ≤0.015 |
| 4 | L2.2.1 | Peru | Positive | 28 | Male | *fgd1* T960C | 2HRZE, 4RH | Unknown | ≤0.015 |
| 5 | L4.3.2 | Peru | Negative | 21 | Male | *fbiC* C1161T | 2HRZE, 4RH | Cured | ≤0.015 |
| 6 | L4.1.2.1 | Peru | Positive | 45 | Male | *fgd1* Lys270Met | 2HRZE, 4RH | Unknown | 0.03 |
| 7 | L4.1.2.1 | Peru | Positive | 36 | Male | *fbiC* G-11A<br>*fgd1* Lys270Met | 2HRZE, 4RH | Unknown | ≤0.015 |
| 8 | L4.1.2 | South Africa | Negative | 57 | Female | *fbiA* Ile208Val | 2HRZE, 4RH | Cured | ≤0.015 |
| 9 | L2.2.1 | Thailand | Unknown | 76 | Male | *fgd1* T960C<br>*ddn* 85-87del (Tyr29del) | 2HRZE, 4RH | Died | >8 |
| 10 | L1.1.1 | Thailand | Negative | 42 | Male | *fbiC* Ala416Val, Trp678Gly<br>*fgd1* Arg64Ser, T960C | 2HRZE, 4RH | Unknown | 0.03 |

299 Abbreviations: MIC, minimal inhibitory concentration; No, number; L, lineage; wt, wild-type; H, isoniazid; R, rifampicin; Z, pyrazinamide; E, ethambutol.

In summary, in the subset of ten isolates with polymorphisms in the six targeted genes, six had no elevated MIC in the microdultion, while four isolates had (Table 1). In line with previous studies, we found that Lys270Met in *fgd1* is a natural polymorphism characteristic of *Mtb* lineage *4.1.2.1*, which may (patient 1 and 6) or may not (patient 7) lead to an increased delamanid MIC (19, 34, 35). All 16 strains of lineage *4.1.2.1* showed this lineage-specific marker (Supplementary Table 1). Furthermore, T960C (*fgd1*) is a synonymous substitution and was found in three other patient isolates which expectedly did not have a critical MIC. The increase in the delamanid MIC in the isolate of patient 9 was due to the deletion in *ddn* (7). Our results thus suggest that Tyr29del is a natural polymorphism leading to an increased delamanid MIC. Our study was too small to estimate the prevalence of strains that are naturally resistant to delamanid. Lee et al. 2020, screened 14,876 *Mtb* strains and found two strains with Tyr29del, for a prevalence of 0.013% (36). However, in their study, only the *ddn* gene was screened and the prevalence of natural resistance could, therefore, be higher.

In conclusion, we confirm that mutations in F_420_ genes can confer an elevated delamanid MIC (13, 19). Whether our findings also apply to the related drug pretomanid should be investigated in future studies. The occurrence of clinical *Mtb* isolates with naturally elevated MICs to delamanid from previously untreated patients calls for careful drug susceptibility testing (DST) prior to delamanid treatment (5, 36). However, access to DST is limited in high burden countries. This dilemma highlights the conflict between making new drugs available in high-burden countries and avoiding spread of drug-resistant strains.

## Data Availability

WGS data from patients Mtb strains shown in Table 1 have been submitted to the NCBI (PRJNA300846; Supplementary Table 1).

## Data availability

WGS data from patients *Mtb* strains shown in Table 1 have been submitted to the NCBI (PRJNA300846; Supplementary Table 1).

## ACKNOWLEDGMENTS

We thank all sites that participated, patients whose data were used in this study and Marie Ballif for contributing to the data collection and for critically reading the manuscript. Calculations were performed on UBELIX (http://www.id.unibe.ch/hpc), the HPC cluster at the University of Bern.

## ROLE OF FUNDING SOURCE

This research was supported by the Swiss National Foundation (project grant numbers 153442, 310030_166687, 310030_188888, IZRJZ3_164171, IZLSZ3_170834 and CRSII5_177163). The International Epidemiology Databases to Evaluate AIDS (IeDEA) is supported by the U.S. National Institutes of Health’s, National Institute of Allergy and Infectious Diseases, the Eunice Kennedy Shriver National Institute of Child Health and Human Development, the National Cancer Institute, the National Institute of Mental Health, the National Institute on Drug Abuse, the National Heart, Lung, and Blood Institute, the National Institute on Alcohol Abuse and Alcoholism, the National Institute of Diabetes and Digestive and Kidney Diseases, the Fogarty International Center, and the National Library of Medicine: Asia-Pacific, U01AI069907; CCASAnet, U01AI069923; Central Africa, U01AI096299; East Africa, U01AI069911; NA-ACCORD, U01AI069918; Southern Africa, U01AI069924; West Africa, U01AI069919. RJW receives support from the Francis Crick Institute, which is funded by UKRI, CRUK, and Wellcome (FC0010218, 104803, 203135). This work is solely the responsibility of the authors and does not necessarily represent the official views of any of the institutions mentioned above. This manuscript is not peer-reviewed.

## CONFLICT OF INTEREST

Authors have nothing to disclose.

## Notes

### Competing Interest Statement

The authors have declared no competing interest.

### Author Declarations

The Cantonal Ethics Committee in Bern, Switzerland, and local institutional review boards approved the study. Written informed consent was obtained at all locations, except in South Africa, where consent was not required for archived samples.

